# Rapid direct-from-urine growth detection to triage culture-negative urine: a prospective diagnostic accuracy study of a two-tube nitrate-reduction and bromothymol blue assay (SIBURI/SIBURI+)

**DOI:** 10.64898/2026.10.29.26366819

**Authors:** Roshan Ratnakar Naik, Lakshmi Swaroopa Naik, Pradnya Naik, Rakshat Eknath Paryekar, Anita Sandhya Estibeiro

## Abstract

**Objectives:** Urine culture takes 24–72 h to report, and most specimens are culture-negative. We evaluated whether a 6-h direct-from-urine growth-detection assay can identify culture-negative specimens.

**Methods:** In a prospective single-centre study at Goa Medical College, 145 consecutive urine specimens from patients with suspected urinary tract infection (UTI) were tested in parallel. Urine (50 µL) was added to SIBURI (nitrate-reduction broth, developed with Griess reagent) and SIBURI+ (Luria-Bertani broth with bromothymol blue) and incubated for 6 h at 37 °C. Any visible colour change against an uninoculated blank was scored growth-positive. The reference standard was quantitative culture (≥10□ CFU/mL of a single organism). Twenty-five specimens reported as contaminated, “SCC” or with _≥_3 organisms were excluded. Accuracy was estimated with exact 95% confidence intervals. A cell-free nitrite series and ATCC *Escherichia coli* and *Klebsiella pneumoniae* dilution series characterised the colour response.

**Results:** Of 120 analysed specimens, 41 (34.2%) were culture-positive. Sensitivity was 92.7% (95% CI 80.1–98.5), specificity 92.4% (84.2–97.2), negative predictive value (NPV) 96.1% (88.9–99.2) and positive predictive value 86.4% (72.6–94.8). SIBURI alone detected 29/41 culture-positive specimens (70.7%); nine (enterococci, streptococcus, *Acinetobacter, Candida* and two unspecified) were detected only by SIBURI+. Classifying the excluded specimens as culture-positive gave a sensitivity of 93.9% and an NPV of 94.8%; classifying them as culture-negative lowered specificity to 71.2%. The colour response was biphasic: pink to red from 10 µM to 1 mM nitrite, and yellow above 1.5 mM.

**Conclusions:** In this single-centre study, a two-tube 6-h assay showed good accuracy against culture, but the lower confidence limits (sensitivity 80.1%, NPV 88.9%) do not yet support omitting culture after a negative result. Positive results need culture confirmation, and larger multicentre validation is required.

## 1. Introduction

Urinary tract infections (UTIs) are among the most common bacterial infections in community and hospital practice, and roughly half of all women have at least one episode in their lifetime [1,2]. *Escherichia coli* causes most cases, followed by *Klebsiella pneumoniae, Proteus mirabilis, Enterococcus faecalis* and *Staphylococcus saprophyticus* [2]. Timely, accurate diagnosis supports targeted treatment and antimicrobial stewardship [3].

At the point of care, diagnosis relies on urine dipstick analysis, a rapid (about 2 min) qualitative test for nitrite and leukocyte esterase. The nitrite reaction depends on bacterial reduction of urinary nitrate. In a meta-analysis, nitrite had a pooled sensitivity of about 45–60% and a specificity of about 85–98% [4], so a negative result cannot exclude infection, and the dipstick gives no information on the organism. When symptoms persist, urine culture is recommended. Culture is the reference standard, with a count of ≥10□ CFU/mL defining significant bacteriuria [5,6]. It is slow, however: isolation on differential media (about 24 h), biochemical identification (about 24 h) and susceptibility testing (about 24 h) together delay a definitive result by 48–72 h.

Culture also burdens laboratories, because a large proportion of specimens submitted for culture are reported as no significant growth [7]. Each of these still needs 24–48 h of incubation before the negative result can be issued, consuming laboratory capacity and delaying reassurance, or the search for an alternative diagnosis, for patients without bacterial UTI. A same-day presumptive test that identifies likely culture-negative specimens, used as a triage step ahead of full culture and not as a replacement for it, could reduce this burden. Rapid point-of-care systems for bacteriuria are being developed, for example a nanofluidic analyser that reported 84.0% sensitivity and 99.4% specificity for species within its specification [8], but such instruments are costly, and a low-cost visual assay would be complementary.

Because *E. coli* and most other Enterobacterales reduce nitrate to nitrite, a growth-based nitrite reaction is an attractive basis for such a test. Enterococci and *S. saprophyticus*, however, generally lack nitrate reductase and are recognised causes of false-negative nitrite results. A nitrate-reduction read-out alone would miss them, so a general growth indicator is also needed.

We describe and evaluate a two-tube assay applied directly to urine without prior culture isolation. SIBURI is a nitrate-reduction broth read with Griess reagent for nitrate-reducing organisms. SIBURI+ is Luria-Bertani (LB) broth with bromothymol blue (BTB) for organisms that do not reduce nitrate. We evaluated the growth-detection function of the assay, namely its ability to separate culture-positive specimens (≥10□ CFU/mL) from culture-negative specimens, against standard urine culture, with a read-out at 6 h. We also characterised the colour chemistry of the assay with nitrite standards and reference strains. The antibiotic-susceptibility function of the kit is outside the scope of this study.

## 2. Materials and methods

### 2.1 Study design and ethics

This was a prospective, single-centre diagnostic accuracy study, reported according to STARD 2015 [9]. It was conducted in the Department of Microbiology, Goa Medical College, Bambolim, Goa, India between May and October 2025. The Institutional Ethics Committee approved the study (GMCIEC/2023/306) with waiver of informed consent.

### 2.2 Specimens and eligibility

Mid-stream clean-catch urine specimens from outpatients and inpatients with clinically suspected UTI, submitted for routine culture, were enrolled consecutively. Specimens with inadequate volume, or in unlabelled, leaking or non-sterile containers, were excluded. All freshly collected urine specimens transported on ice, received until 12 PM in the department of microbiology on the day were streaked on appropriate culture media and SIBURI, SIBURI+ analysis in parallel within one hour of receipt. After culture, specimens reported as contaminated or squamous cell contamination (SCC), or with _≥_3 organism types (mixed growth) were excluded from the accuracy analysis, because they cannot be assigned an unambiguous reference status. A single rule, based on the reported culture result, was applied to all specimens and this criterion was set before analysis. The index-test results for excluded specimens were retained and are reported in Section 3.4. The flow of specimens is shown in Fig. 1.

**Fig. 1.**
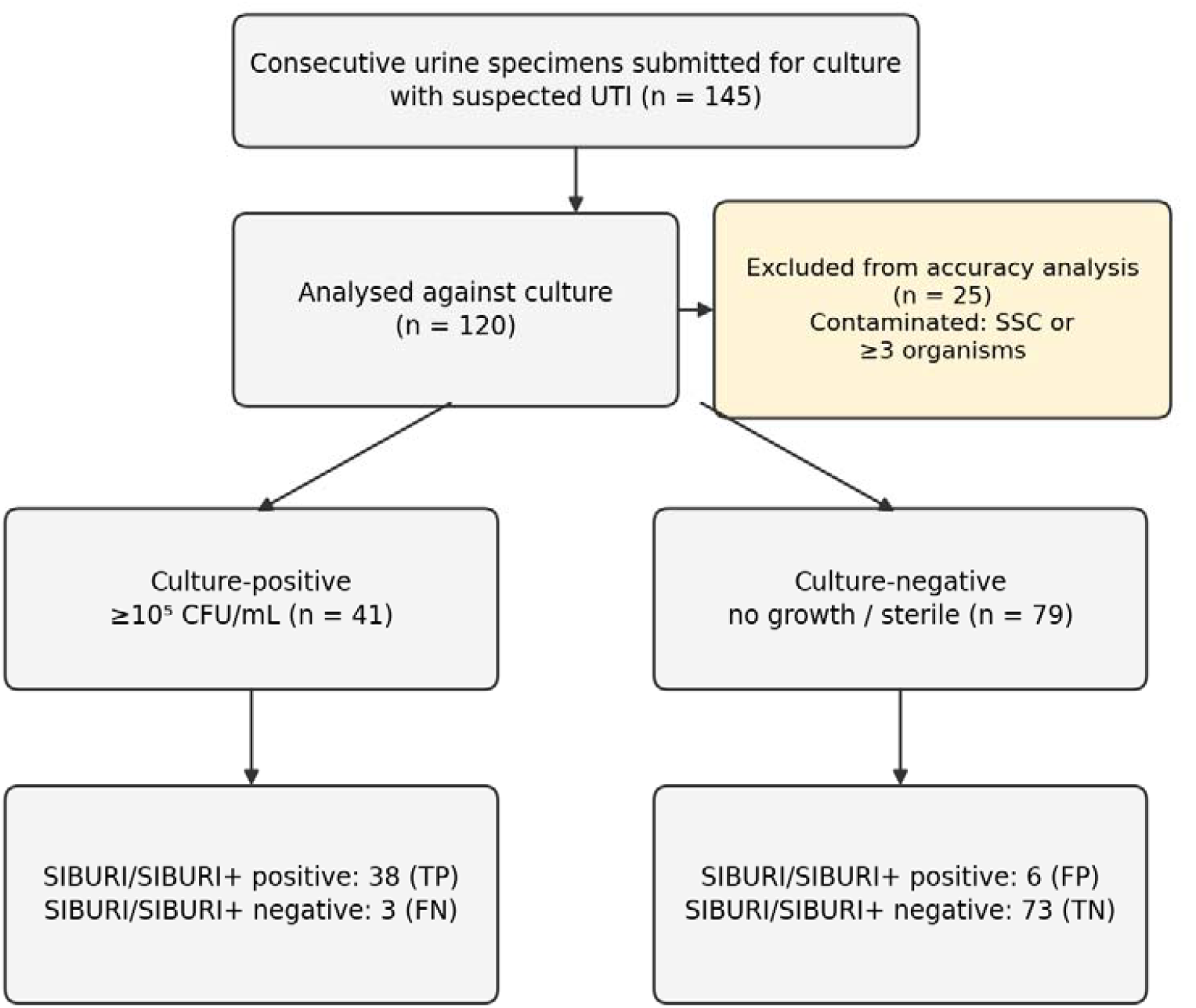
Flow of specimens through the study (STARD diagram). TP, true positive; FN, false negative; FP, false positive; TN, true negative.

### 2.3 Reference standard: urine culture

Urine was inoculated with a calibrated loop (0.001 mL) onto MacConkey agar without crystal violet, with 0.5% bile salt (HiMedia, Mumbai, India), and incubated aerobically at 37 °C for 18–24 h. Growth of ≥10□ CFU/mL of a single organism was classified as culture-positive (significant bacteriuria). No growth and sterile results were classified as culture-negative. Organisms were identified by colony morphology, Gram stain and standard biochemical tests as part of the laboratory’s routine workflow. Only the categorical culture result was recorded, and individual CFU/mL counts were not available for analysis and the culture readers were blinded to the index test.

### 2.4 Index test: SIBURI and SIBURI+

#### SIBURI (nitrate-reduction broth)

The broth contained 0.3 g peptone, 0.2 g HM extract and 0.06 g sodium nitrate in 10 mL distilled water (labeled RISA, patent no. 553621 [10A, 10B]), adjusted to pH 7.5 and autoclaved at 121 °C for 15 min. Bacteria with nitrate reductase reduce the nitrate to nitrite during incubation. Nitrite is detected by adding Griess reagent. The Griess reagent was prepared in a final volume of 100 mL containing 10 mL of 85% orthophosphoric acid (Qualigens, Mumbai, India), 1.0 g of sulfanilic acid (Loba Chemie, Mumbai, India) and 0.1 g of _α_-naphthylamine (Clairofilt India, Mumbai, India). The reagents were dissolved and the volume was adjusted to 100 mL with distilled water, in which nitrite forms a diazonium salt that couples to give a coloured azo dye [11]. The colour ranges from yellowish-orange through pink to deep red, depending on nitrite concentration. The kit also contains parallel antibiotic-containing tubes, which are outside the scope of this study; only the no-antibiotic tube was analysed.

#### SIBURI+ (LB broth with bromothymol blue)

LB broth (10 g tryptone, 5 g yeast extract and 10 g NaCl per litre; (HiMedia, Mumbai, India) was adjusted to pH 7.5 and autoclaved at 121 °C for 15 min. A 0.05% (w/v) BTB stock (Loba Chemie, Mumbai, India) was neutralised to pH 7.0 (green) with sterile 1 M NaOH and filter-sterilised (0.22 µm). For each assay, 9.2 mL of sterile LB was supplemented with 800 µL of BTB stock (final 0.004% w/v). SIBURI+ detects growth of organisms that do not reduce nitrate.

#### Procedure

For each specimen, urine was added to SIBURI and, separately, to SIBURI+ in 1.5 mL polypropylene tubes, to a total volume of 1 mL (50 µL urine plus 950 µL broth, a 1:20 dilution). Tubes were incubated for 6 h at 37 °C in a dry-bath incubator. 25 µL of Griess reagent were added to the SIBURI tube and the colour was read after 1 min. The SIBURI+ colour was read without further additions. An uninoculated broth blank (bluish) and a positive control strain (yellow-green) were included with each batch.

#### Scoring

Each tube was compared visually with an uninoculated blank. Any visible colour change relative to the blank (yellow, orange, pink, red or darker for SIBURI; blue to green or yellow-green for SIBURI+ (Fig. 2). A specimen was index-test positive if either tube was growth-positive and index-test negative if neither was. The colour chart developed from the analytical experiments (Section 2.6) was used to describe colour, not to define the positivity threshold. Two readers blinded to culture results recorded the color change independently.

**Fig. 2.**
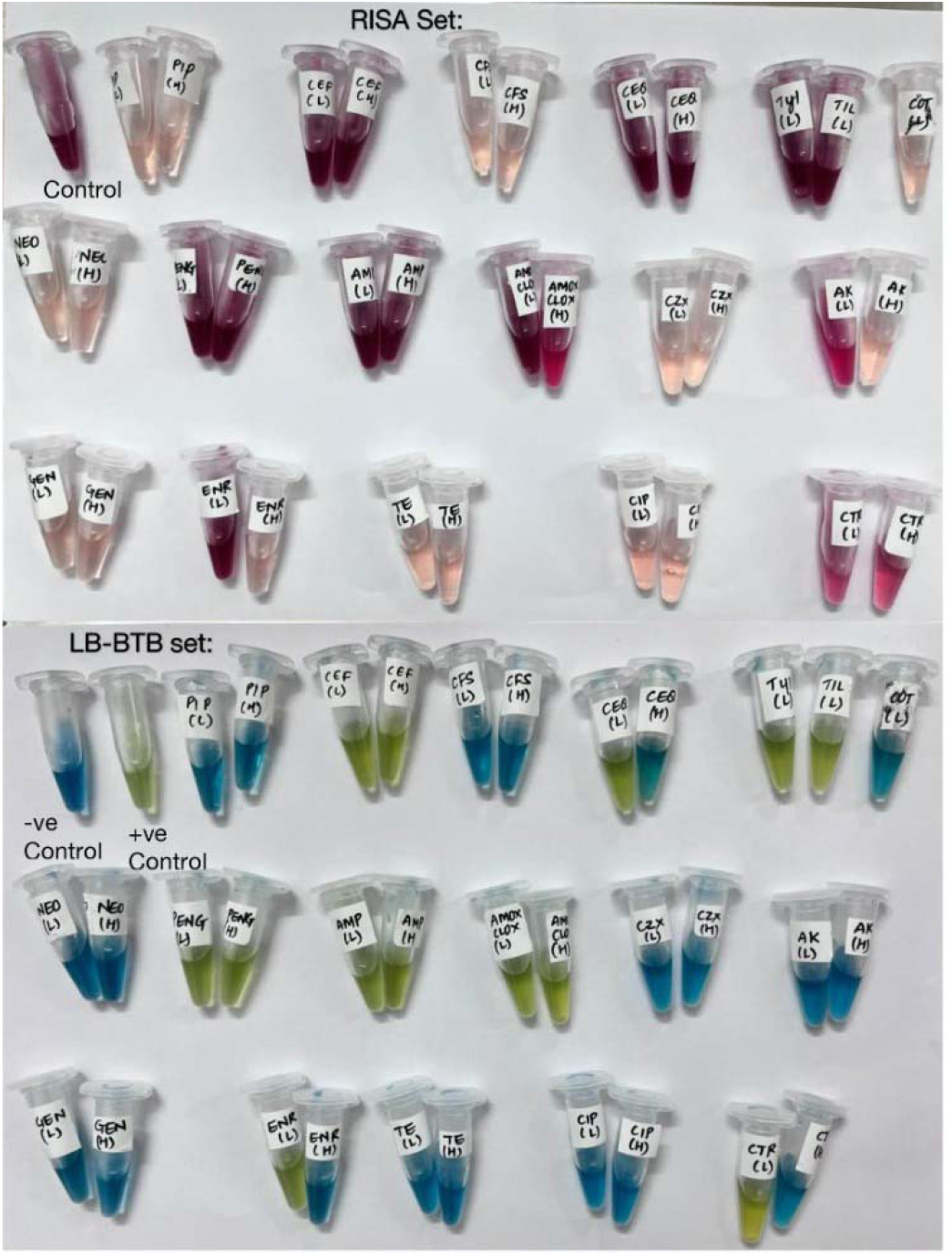
Representative SIBURI (labelled RISA in the photograph; top) and SIBURI+ (LB with bromothymol blue, LB-BTB; bottom) tubes after 6 h at 37 °C. The first tube of each set is the control; in SIBURI+, the uninoculated control (−ve, blue) and the inoculated control (+ve, yellow-green) are shown. The remaining tube pairs contain antibiotics at low (L) and high (H) concentrations; these susceptibility tubes are outside the scope of this study, and only the no-antibiotic control tubes were analysed.

### 2.5 Sample size

Sample size was estimated for a diagnostic accuracy study with the Buderer formula [12,13]: n(Se) = Z^2^·Se(1−Se)/(d^2^·Prev) and n(Sp) = Z^2^·Sp(1−Sp)/(d^2^·(1−Prev)), where Z = 1.96, Se and Sp are the anticipated sensitivity and specificity, d is the desired precision and Prev is the prevalence of culture-confirmed UTI. We assumed Se = 90% and Sp = 85% [14-16], d = 0.08 (95% confidence interval half-width of 8 percentage points) and Prev = 45%, the mid-point of the 42–48% reported among suspected cases in Indian tertiary care [17-19]. This gave n(Se) = 121 and n(Sp) = 140. Specificity set the requirement at 140, and an allowance of 5% for exclusions gave 147 specimens (about 66 culture-positive and 81 culture-negative expected). This is a precision-based calculation and not a power calculation for a hypothesis test. In terms of numbers of each reference class, the calculation required about 55 culture-positive and 77 culture-negative specimens.

### 2.6 Analytical characterisation of the colour response

#### Sodium nitrite series

To characterise the colour chemistry independently of bacterial growth, a 1 M sodium nitrite stock (0.69 g in 10 mL distilled water; Loba Chemie) was serially diluted in uninoculated SIBURI broth to 0.1, 1, 10, 30, 50, 70, 100 and 500 µM and 1, 1.5, 2, 2.5, 5, 10, 30, 50, 70, 100 and 500 mM. Each concentration was prepared in duplicate (1 mL) with a reagent blank (1 mL broth). After 10 min at room temperature, 20 µL of Griess reagent was added and mixed. The colour was recorded visually and photographed (Fig. 2). Absorbance was scanned using scan-mode run between 340 and 700 nm against the reagent blank (Kanad 8MPC UV-visible spectrophotometer, Kanad Vidyut, Pune, India), and values at 450 and 545 nm are reported.

#### Reference-strain dilution series

*Escherichia coli* ATCC 25922 and *Klebsiella pneumoniae* ATCC 13883 were grown overnight at 37 °C in sterile SIBURI broth and adjusted to 0.5 McFarland (about 1.5 × 10□ CFU/mL; A630 _≈_ 0.09 against sterile broth), labelled 10□. Ten-fold serial dilutions (100 µL into 900 µL broth) gave the 10□ to 10□ tubes. The remaining 900 µL of each dilution was incubated at 37 °C for 6 h, 25 µL Griess reagent was added and the colour was read after 1 min. Because clinical specimens were diluted 1:20 in broth, a urine count of 10□ CFU/mL corresponds to about 5 × 10^3^ CFU/mL at the start of incubation.

### 2.7 Statistical analysis

Index-test results were cross-tabulated against culture. Sensitivity, specificity, positive and negative predictive values (PPV, NPV), accuracy and likelihood ratios were calculated, with exact (Clopper–Pearson) 95% confidence intervals [19]. The contribution of SIBURI+ was assessed from the specimens that were positive only on SIBURI+, using an exact sign test. Two sensitivity analyses examined the effect of the exclusions: classifying all excluded specimens as culture-negative, and classifying all of them as culture-positive. The analytical experiments were descriptive. Statistical analysis and calculations were performed using Python 3.12.11 with pandas library v3.0.6, SciPy v1.16.3 and NumPy v2.1.3.

## 3. Results

### 3.1 Study population

Of 145 specimens enrolled, 25 (17.2%) were excluded from the accuracy analysis under a single rule: 23 were reported as SCC and 2 as three organism types (Fig. 1). A total of 120 specimens were analysed, of which 41 (34.2%; 95% CI 25.8–43.4%) were culture-positive and 79 were culture-negative. The study population comprised both male and female patients, aged 18–75 years, including both inpatients and outpatients.

### 3.2 Diagnostic accuracy

Against culture, the combined SIBURI/SIBURI+ read-out at 6 h was positive in 38 of 41 culture-positive specimens and negative in 73 of 79 culture-negative specimens (Table 1). Sensitivity was 92.7% (95% CI 80.1–98.5%), specificity 92.4% (84.2–97.2%), NPV 96.1% (88.9–99.2%), PPV 86.4% (72.6–94.8%) and accuracy 92.5% (86.2–96.5%). The positive likelihood ratio was 12.2 and the negative likelihood ratio was 0.079. Overall, 76 of 120 specimens (63.3%) were index-negative, and 73 of these (96.1%) were culture-negative.

**Table 1.**
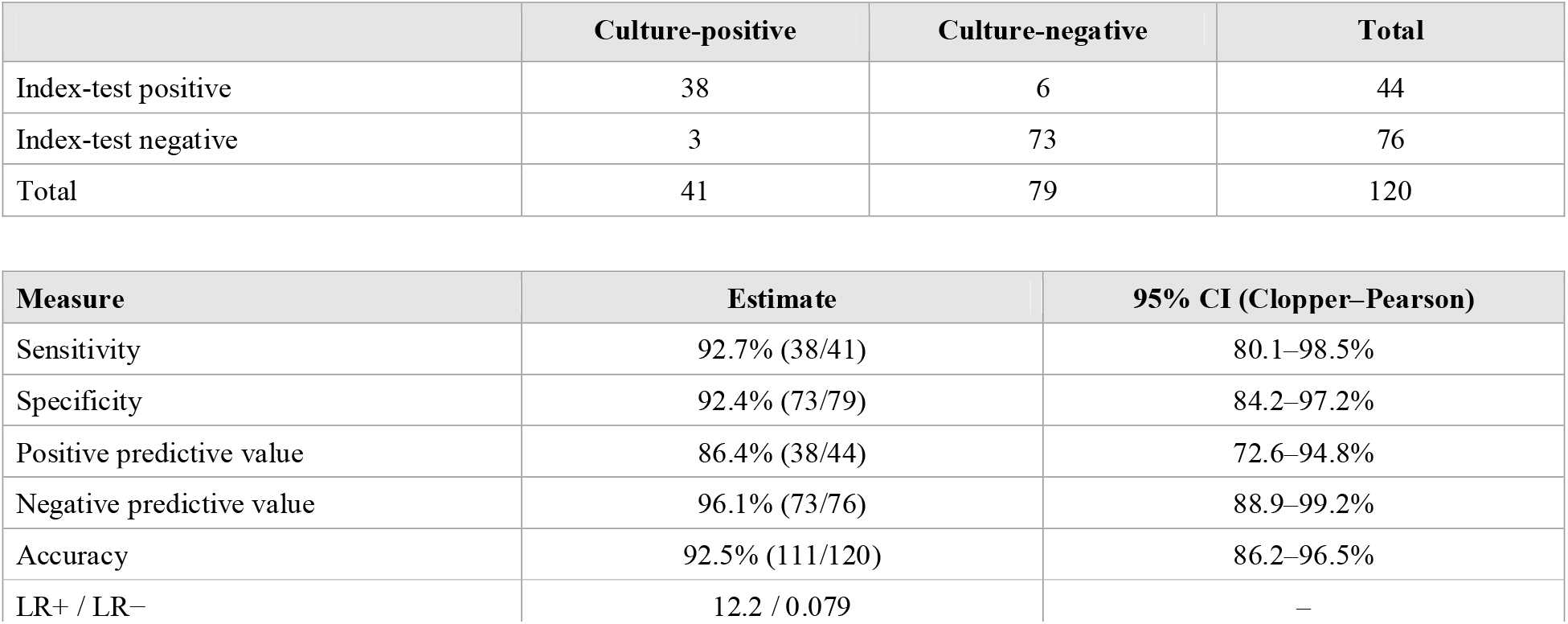
Accuracy of SIBURI/SIBURI+ against urine culture (n = 120)

The three false negatives were culture-positive specimens with no colour change in either tube: one *Enterococcus*, one *Pseudomonas aeruginosa* and one with no organism recorded. Of the six false positives, five were described as faint yellowish, light pink, pink or orange and one as dark red. Three of the six occurred among specimens recorded as sterile.

### 3.3 Contribution of each tube and organism pattern

Nine of the 38 true positives (23.7%) showed no colour change in SIBURI and were detected only by SIBURI+. These were four *Enterococcus* isolates, one *Streptococcus*, one *Acinetobacter*, one non-*albicans Candida* and two culture-positive specimens with no recorded organism (Table 2). Had SIBURI alone been used, sensitivity would have been 70.7% (29/41; 95% CI 54.5–83.9%) with unchanged specificity, and NPV 85.9% (73/85). All nine discordant results favoured the combined read-out (exact sign test, *P* = 0.004). All *E. coli* and *Klebsiella* specimens were detected by SIBURI; one of two *Pseudomonas aeruginosa* specimens and one of five enterococcal specimens were missed by both tubes.

**Table 2.** Index-test positivity by culture result among culture-positive specimens (n = 41)

| **Culture result** | **n** | **Detected** | **Detected only by SIBURI+** |
|---|---|---|---|
| *Escherichia coli* | 4 | 4 | 0 |
| *Klebsiella pneumoniae* | 2 | 2 | 0 |
| *Pseudomonas aeruginosa* | 2 | 1 | 0 |
| *Enterococcus* spp. | 5 | 4 | 4 |
| Other (Gram-positive, *Streptococcus*, *Acinetobacter*) | 4 | 4 | 2 |
| Polymicrobial (two organisms) | 5 | 5 | 0 |
| Yeast-containing | 3 | 3 | 1 |
| Organism not recorded | 16 | 15 | 2 |
| Total | 41 | 38 | 9 |

### 3.4 Excluded specimens and sensitivity analyses

Among the 25 excluded specimens, 24 (96.0%) were index-test positive, consistent with growth of mixed or commensal flora in the broths. To bound the effect of the exclusions, we re-analysed all 145 specimens with the excluded specimens classed first as culture-negative and then as culture-positive (Table 3).

**Table 3.** Sensitivity analyses.

| **Analysis** | **n** | **Sensitivity (95% CI)** | **Specificity (95% CI)** | **NPV (95% CI)** |
|---|---|---|---|---|
| Primary analysis | 120 | 92.7% (80.1–98.5) | 92.4% (84.2–97.2) | 96.1% (88.9–99.2) |
| Excluded specimens counted as culture-negative | 145 | 92.7% (80.1–98.5) | 71.2% (61.4–79.6) | 96.1% (89.0–99.2) |
| Excluded specimens counted as culture-positive | 145 | 93.9% (85.2–98.3) | 92.4% (84.2–97.2) | 94.8% (87.2–98.6) |

### 3.5 Analytical characterisation: sodium nitrite series

In the cell-free nitrite series measured using scan-mode run (Fig. 3, Table 5), tubes at 0.1 and 1 µM were colourless to pale yellow. Pink colour appeared from 10 µM and deepened through 30–70 µM to a strong red at 100 µM, 500 µM and 1 mM. At 1.5 mM the tubes were orange-yellow, and from 2 mM upward they were yellow (paler at 500 mM); the change from red to yellow between 1 mM and 1.5 mM was abrupt. Absorbance at 545 nm followed the pink/red colour only up to this point (A545 0.0065 at 0.1 µM, rising to 1.997 at 500 µM and 1.818 at 1 mM), then fell about nine-fold to 0.195 at 1.5 mM and stayed at 0.06–0.28 up to 500 mM (Fig. 4). Absorbance at 450 nm, by contrast, remained high (1.13–2.22) from 500 µM to 500 mM, in keeping with the yellow appearance.

**Table 5.** Sodium nitrite standard series: visual colour and scan-mode absorbance.

| NaNO□ | Visual colour (Fig. 3) | A450 | A545 | A450 : A545 |
|---|---|---|---|---|
| 0.1 µM | Colourless / pale yellow | 0.009 | 0.0065 | – |
| 1 µM | Colourless / pale yellow | 0.016 | 0.0145 | – |
| 10 µM | Pale peach | 0.063 | 0.124 | 0.51 |
| 30 µM | Salmon pink | 0.167 | 0.338 | 0.49 |
| 50 µM | Pink-red | 0.277 | 0.617 | 0.45 |
| 70 µM | Pink-red | 0.382 | 0.767 | 0.50 |
| 100 µM | Red | 0.536 | 1.114 | 0.48 |
| 500 µM | Deep red | 1.744 | 1.997 | 0.87 |
| 1 mM | Deep red | 2.216 | 1.818 | 1.22 |
| 1.5 mM | Orange-yellow | 1.562 | 0.195 | 8.01 |
| 2 mM | Yellow | 1.521 | 0.177 | 8.59 |
| 2.5 mM | Yellow | 1.532 | 0.171 | 8.96 |
| 5 mM | Golden yellow-orange | 1.872 | 0.276 | 6.78 |
| 10 mM | Yellow | 1.619 | 0.195 | 8.32 |
| 30 mM | Yellow | 1.562 | 0.164 | 9.52 |
| 50 mM | Yellow | 1.591 | 0.147 | 10.86 |
| 70 mM | Yellow | 1.580 | 0.141 | 11.21 |
| 100 mM | Yellow | 1.615 | 0.135 | 11.96 |
| 500 mM | Pale yellow | 1.129 | 0.060 | 18.97 |

Colour descriptions were assigned from the photograph in Fig. 3 and were not independently rated. The A450 : A545 ratio is descriptive only and is omitted at 0.1 and 1 µM, where both readings were near the blank.

**Fig. 3.**
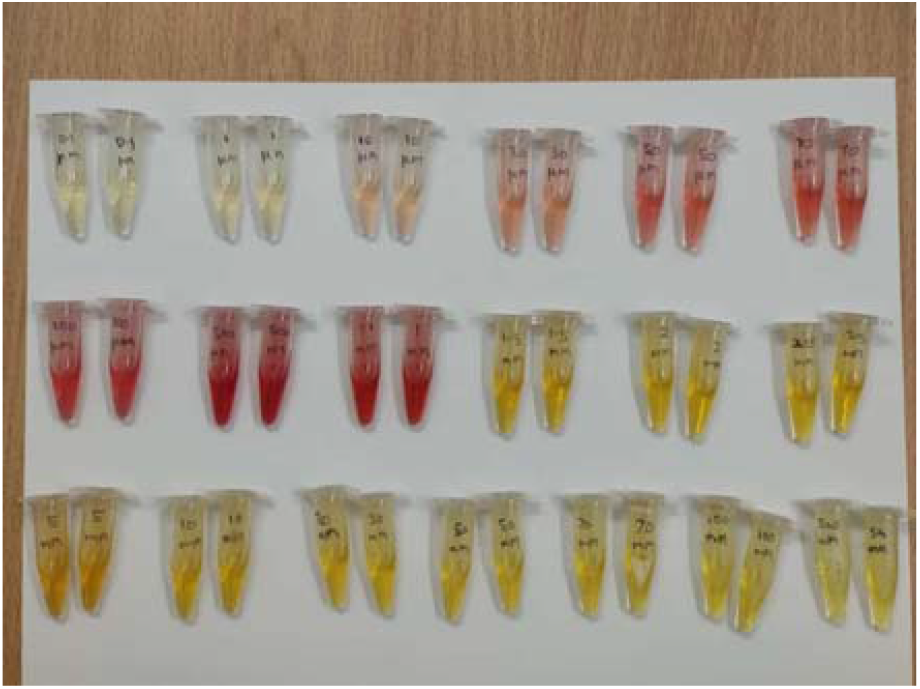
Cell-free sodium nitrite standard series (duplicate tubes; 0.1, 1, 10, 30, 50, 70, 100, 500 µM and 1, 1.5, 2, 2.5, 5, 10, 30, 50, 70, 100, 500 mM) in SIBURI broth, photographed 10 min after addition of Griess reagent. Colour deepens from colourless through pink to red up to 1 mM and then changes abruptly to orange and yellow at higher concentrations.

**Fig. 4.**
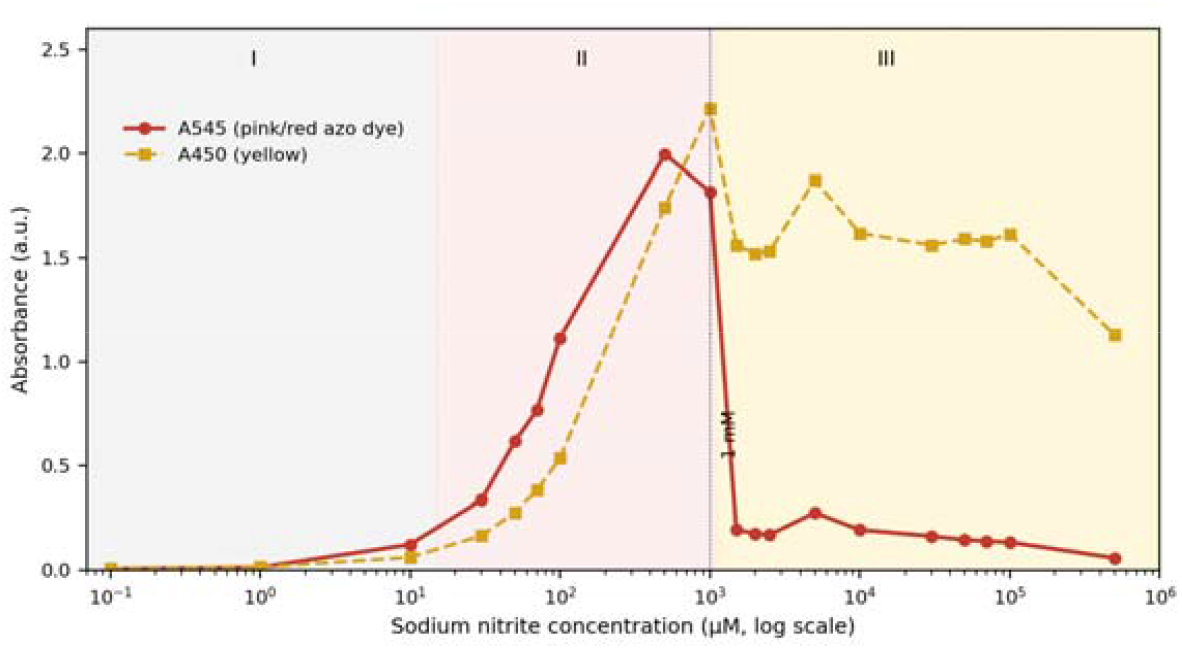
Absorbance of the sodium nitrite standard series at 545 nm (red) and 450 nm (yellow) from the scan-mode measurements (log concentration axis). Regions: I, low nitrite (0.1–10 µM), little colour; II, increasing response (10 µM to about 1 mM), colourless to pink/red; III, high nitrite (>1 mM), colour shifts to orange/yellow and A545 falls.

A single-wavelength reading at 545 nm therefore cannot separate a low-nitrite tube from a very-high-nitrite tube: the yellow 5 mM tube read A545 0.276, close to the pale-pink 30 µM tube at 0.338. The hue of the tube, not its 545 nm absorbance, carries the information. Separate runs showed the same qualitative pattern, but absolute A545 values differed between runs (for example, 100 µM read 1.33, 0.656 and 1.114 in three runs), which argues against using absolute absorbance as the read-out.

### 3.6 Analytical characterisation: CFU/mL colour response

Both ATCC reference strains showed the same biphasic colour response across the 10□–10□ dilution series (Fig. 5, Table 6): strong yellow at 10□–10□, red at 10□, bright pink at 10□, and minimal or no colour at ≤10^3^ CFU/mL. This is the pattern expected if nitrite accumulates with increasing bacterial density: yellow tubes lie beyond the pink/red working range, red and pink tubes lie within it, and colourless tubes lie below the visual detection limit. The concordance is qualitative only, because nitrite was not measured in the bacterial tubes and the CFU/mL values are nominal dilution labels.

**Table 6.**
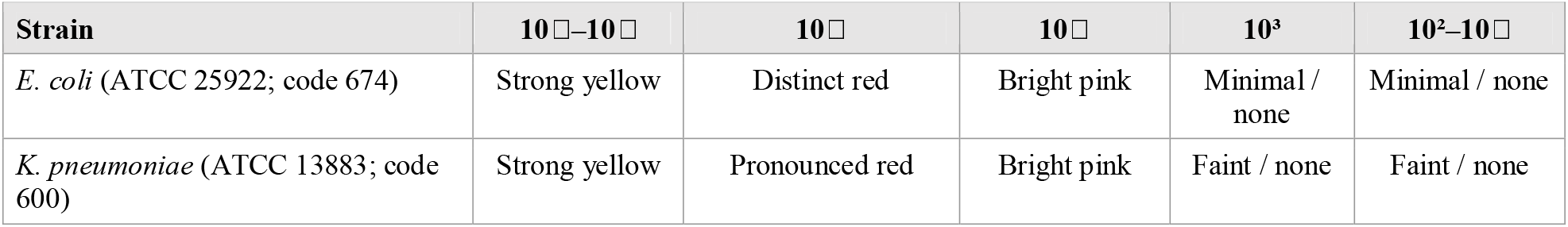
Colour response in SIBURI after 6 h across ten-fold dilutions (nominal CFU/mL)

**Fig. 5.**
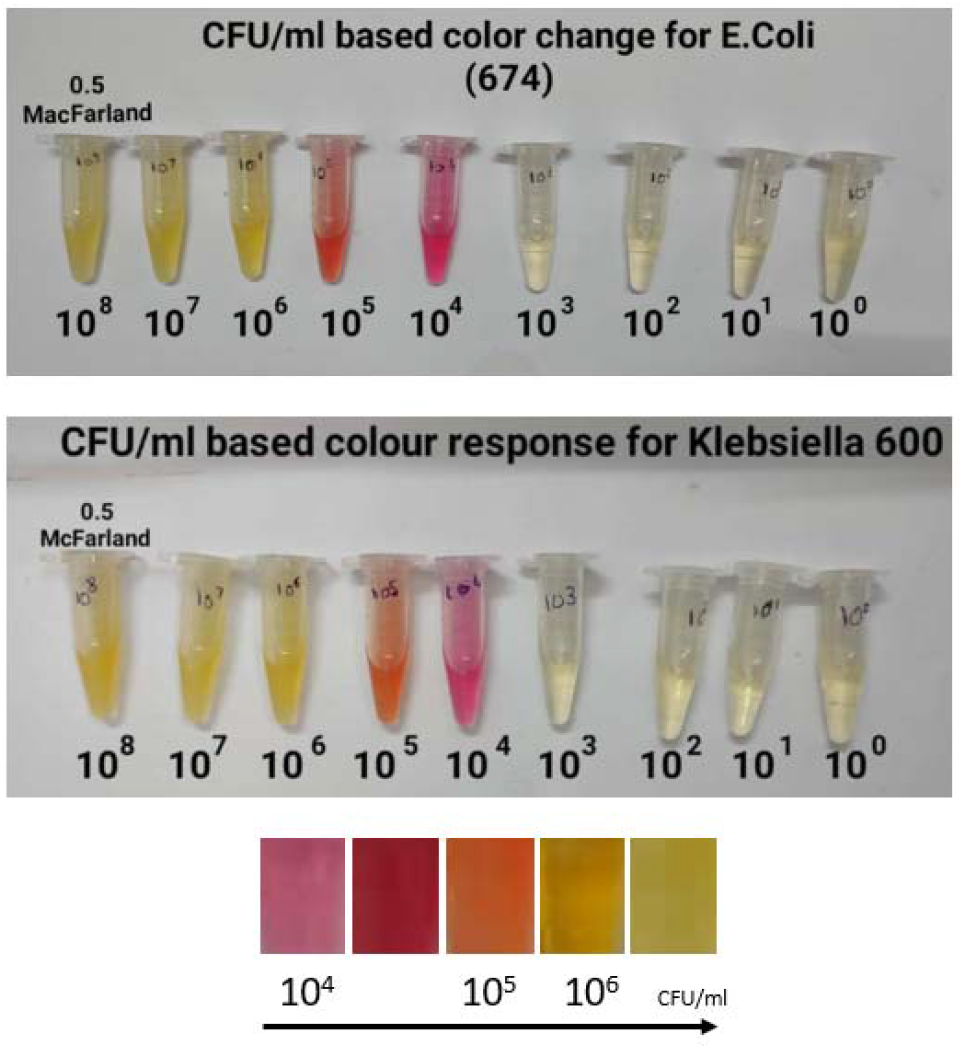
Colour response of the SIBURI–Griess reagent system after 6 h across ten-fold dilutions (10□–10□ CFU/mL, nominal; 10□ = 0.5 McFarland) of *E. coli* (ATCC 25922, laboratory code 674, top) and *Klebsiella* (ATCC 13883, laboratory code 600, bottom). The kit colour chart with approximate CFU/mL labels is shown at the bottom.

## 4. Discussion

### 4.1 Principal findings

In this single-centre study, a two-tube, 6-h, direct-from-urine assay detected 38 of 41 culture-positive specimens and correctly classified 73 of 79 culture-negative specimens. For a triage test, whose purpose is to identify specimens unlikely to need further work-up, the NPV is the operationally important metric. The NPV was 96.1%, but the lower confidence limit was 88.9%, and the NPV depends on prevalence (34.2% here). A same-day negative result for about 63% of specimens would substantially reduce the culture workload if confirmed, but three of the 76 index-negative specimens were culture-positive, and the interval does not yet support omitting culture after a negative result, particularly for high-risk patients. The PPV of 86.4% means that positive results must still be confirmed by culture and cannot be reported as UTI.

### 4.2 Complementary roles of the two tubes

The two tubes serve different organism groups. The nitrate-reduction tube detected the Enterobacterales, whereas nine culture-positive specimens, including four of the five enterococcal specimens, were detected only by the bromothymol blue tube. Without SIBURI+, sensitivity would have been 70.7%. This is consistent with the limitation of nitrite-based methods for organisms that lack nitrate reductase [4]. Because SIBURI+ reports growth of any organism, it also cannot distinguish a pathogen from a contaminant, which is relevant to the false positives discussed below.

### 4.3 Comparison with other rapid tests

The nitrite pad of the dipstick shows a pooled sensitivity of about 45–60% [4], well below the 92.7% observed here, but the comparison is indirect, because the dipstick is read within minutes of voiding and our assay needs a 6-h incubation; it trades speed for sensitivity. A nanofluidic point-of-care analyser reported a sensitivity of 84.0% (89/106) and a specificity of 99.4% (155/156) in 278 women with suspected uncomplicated UTI [8]. Our sensitivity is numerically higher and our specificity lower. The populations, reference methods and organism scopes differ, and our sample is far smaller, so no head-to-head conclusion can be drawn. A direct comparison with a nitrite dipstick recorded on the same specimens is the most useful next step.

### 4.4 False positives, contaminated specimens and reader effects

Six specimens were false positives. Five were described with faint or borderline colours, consistent with reader subjectivity on a continuous colour scale. A positive 6-h read-out reflects growth, not a count of ≥10□ CFU/mL, so specimens with lower counts that grow within 6 h would be scored positive and classified as false positives; because individual CFU/mL values were not recorded, we could not test this. Pre-existing nitrite in the specimen, or bacterial growth in the container before testing, could also contribute. The three false negatives, and three of the six false positives were among specimens recorded as sterile, a group that merits review of collection and handling. The false negatives were one enterococcal specimen (no colour change in either tube), one *P. aeruginosa* specimen and one specimen with no organism recorded. *P. aeruginosa* may reduce nitrite further, so that it does not accumulate [20], but this is speculative from a single specimen, and the other *P. aeruginosa* specimen was detected.

Nearly all contaminated or SCC specimens (24 of 25; 96.0%) were index-positive. This is expected for any growth-based test, since commensal flora will grow in non-selective broth, and it means the assay should not be interpreted on specimens that are visibly contaminated or from poorly collected samples. The sensitivity analyses show how much the conclusions depend on how these specimens are classified: if they were all culture-negative, specificity would fall to 71.2%, and if all culture-positive, sensitivity would be 93.9% and NPV 94.8%. This is the main source of uncertainty for the primary estimates and is a consequence of excluding specimens after the culture result was known.

### 4.5 The biphasic colour response

The nitrite series (Figs. 3 and 4) explains the pattern seen with reference strains (Fig. 5): colour intensity peaks at about 10□–10□ CFU/mL and gives way to yellow at 10□–10□ CFU/mL, as in cell-free nitrite at 1.5 mM and above. The high-density yellow therefore appears to be a property of the detection chemistry at high nitrite concentration, and not evidence that bacteria have stopped reducing nitrate. The chemical basis of the colour loss was not investigated. Three practical consequences follow. First, a yellow tube is a positive result with a high bacterial load, and only a blank-like tube is negative. Second, colour cannot be used to back-calculate CFU/mL without knowing which side of the peak a tube lies on. Third, because A545 does not rise beyond about 1 mM nitrite, a single-wavelength colorimeter would score a heavily positive yellow tube as weakly positive or negative, so visual reading against the chart is essential. The A450 : A545 ratio (about 0.45–0.51 for 10–100 µM and at least 6.8 from 1.5 mM) suggests that two-wavelength or hue-based reading could be evaluated in future, but this was not tested on bacterial cultures or clinical specimens. In addition, the calibration densities are those at the start of incubation, and clinical specimens were diluted 1:20, so the colour–CFU/mL relationship has not been validated against clinical counts. The highest nitrite concentrations in the standard series (above about 70 mM, the maximum the broth nitrate could generate) are also beyond what bacteria could produce in this medium.

### 4.6 Limitations

This was a small, single-centre study and confidence intervals are wide. The 41 culture-positive specimens were fewer than the 55 required by the sample-size calculation, and the observed prevalence (34.2%) was lower than the 45% assumed; 17.2% of specimens were excluded, against 5% allowed. The sensitivity estimate therefore has a lower confidence limit of 80.1%, and the results need confirmation in a larger cohort. Exclusion of contaminated and SCC specimens after culture may inflate accuracy (see Section 4.4). The reference standard was culture on a single medium, without blood agar or CLED, so fastidious organisms may have been missed; CFU/mL counts and species-level identification were not recorded for many positives (16 of 41 had no organism recorded); and culture readers and index-test readers were blinded. The kit’s control tube is read alongside antibiotic-containing tubes, so the growth call is not fully independent of the susceptibility component, which should be addressed in a dedicated control-tube-only protocol. The calibration and nitrite experiments used Griess reagent with a 10-min development, whereas the kit uses same Griess reagent read at 1 min, and nitrite standards were prepared in broth and not in urine.

## 5. Conclusions

A two-tube, 6-h, direct-from-urine assay showed a sensitivity of 92.7% and an NPV of 96.1% against culture in 120 analysed specimens, with a specificity of 92.4%. The nitrate-reduction tube and the bromothymol blue tube detected complementary organism groups. The colour response is biphasic, so yellow indicates a high bacterial load and the assay must be read visually. These results support further evaluation of the assay as a triage step before culture, with positives confirmed by culture. Larger, multicentre validation with blinded readers, recorded CFU/mL counts and a nitrite-dipstick comparator is needed before routine use.

## Data Availability

All data produced in the present study are available upon reasonable request to the authors

## Declarations

### Ethics approval

Institutional Ethics Committee, Goa Medical College (GMCIEC/2023/306).CTRI/2024/04/065556.

### Data availability

The row-level dataset can be supplied as supplementary material.

### Author contributions (CRediT)

Roshan Naik and Lakshmi Swaroopa Naik conducted the index test. Roshan Naik wrote and edited the manuscript. Rakshat Eknath Paryekar conducted reference urine evaluation and index test. Dr Pradnya Naik and Dr Anita Sandhya Estibeiro contributed towards the manuscript.

